# Exuberant elevation of IP-10, MCP-3 and IL-1ra during SARS-CoV-2 infection is associated with disease severity and fatal outcome

**DOI:** 10.1101/2020.03.02.20029975

**Authors:** Yang Yang, Chenguang Shen, Jinxiu Li, Jing Yuan, Minghui Yang, Fuxiang Wang, Guobao Li, Yanjie Li, Li Xing, Ling Peng, Jinli Wei, Mengli Cao, Haixia Zheng, Weibo Wu, Rongrong Zou, Delin Li, Zhixiang Xu, Haiyan Wang, Mingxia Zhang, Zheng Zhang, Lei Liu, Yingxia Liu

**Affiliations:** Shenzhen Key Laboratory of Pathogen and Immunity, National Clinical Research Center for infectious disease, State Key Discipline of Infectious Disease, Shenzhen Third People’s Hospital, Second Hospital Affiliated to Southern University of Science and Technology, Shenzhen, China

## Abstract

The outbreak of Coronavirus Disease 2019 (COVID-19) caused by the severe acute respiratory syndrome coronavirus 2 (SARS-CoV-2) emerged in Wuhan, December 2019, and continuously poses a serious threat to public health. Our previous study has shown that cytokine storm occurred during SARS-CoV-2 infection, while the detailed role of cytokines in the disease severity and progression remained unclear due to the limited case number. In this study, we examined 48 cytokines in the plasma samples from 53 COVID-19 cases, among whom 34 were severe cases, and the others moderate. Results showed that 14 cytokines were significantly elevated upon admission in COVID-19 cases. Moreover, IP-10, MCP-3, and IL-1ra were significantly higher in severe cases, and highly associated with the PaO_2_/FaO_2_ and Murray score. Furthermore, the three cytokines were independent predictors for the progression of COVID-19, and the combination of IP-10, MCP-3 and IL-1ra showed the biggest area under the curve (AUC) of the receiver-operating characteristics (ROC) calculations. Serial detection of IP-10, MCP-3 and IL-1ra in 14 severe cases showed that the continuous high levels of these cytokines were associated with disease deterioration and fatal outcome. In conclusion, we report biomarkers that closely associated with disease severity and outcome of COVID-19. These findings add to our understanding of the immunopathologic mechanisms of SARS-CoV-2 infection, providing novel therapeutic targets and strategy.

---

Coronaviruses are the largest known viruses with a single positive-sense genome of about 31Kb which could infect a wide range of different species, and mainly infect respiratory and intestinal tract causing a wide range of symptoms ^1^. Six members of coronaviruses that have been found to infect human beings, including HCoV-229E, HCoVOC43, HCoV-NL63, and HCoV-HKU1, Severe Acute Respiratory Syndrome coronavirus (SARS-CoV) and Middle East Respiratory Syndrome coronavirus (MERS-CoV) ^1,2^. Until recently, human infection with the novel severe acute respiratory syndrome coronavirus 2 (SARS-CoV-2) was firstly identified in late December, 2019, in Wuhan, China ^3,4^. As of Feb. 29, 2020, a total of 79824 Coronavirus Disease 2019 (COVID-19) cases with 2870 fatal cases were reported ^5^. Furthermore, another 53 countries have also reported COVID-19 cases, posing a serious threat of public health ^6^. Studies have revealed that penumonia is the most common complications following SARS-CoV-2 infection, and followed by acute respiratory distress syndrome (ARDS) ^7,8^. Inflammation is the body’s first coordinated line of defense against tissue damage caused by either injury or infection, involving activating both the innate and adaptive immune responses ^9^. However, exuberant immune responses following infection have been described as a cytokine storm, associated with excessive levels of proinflammatory cytokines and widespread tissue damage including ARDS ^10^.

Cytokine storm has been found during the infection of influenza viruses ^11-16^ as well as coronavirues ^17-24^, and contributes to acute lung injury and development of ARDS ^2^. Preliminary studies have shown that SARS-CoV-2 infection triggers cytokine storm, and results in the increase of a variety of cytokine/chemokine ^25,26^. However, the detailed role of cytokines in diseases severity and progression remains unknown. In this study, we recruited 53 COVID-19 cases with 34 severe cases and 19 moderate cases defined according to China National Health Commission Guidelines for Diagnosis and Treatment of SARS-CoV-2 infection, and analyzed the plasma cytokine/chemokine profile in detail. Most of the severe cases (73.5%) were ≥ 60 years-old, while the moderate cases were mainly in the group of 16-59 years-old (63.2%) (Table S1). The initial symptoms, co-existing chronic medical conditions, intervals of illness onset to admission and antiviral treatment were similar between the two groups. Complications occurred more frequently in the severe cases, including ARDS, respiratory failure, hepatic and renal insufficiency (Table S1). A complete blood count with differential was assessed for each patient either on the date of hospital admission, or at the earliest time-point and thereafter (Table 1). The percentage of lymphocyte (LYM) and neutrophil (NEU) as well as the CD4 and CD8 counts were significantly lower in the severe cases, indicating a possible dysfunction of immune responses.

**Table 1.**
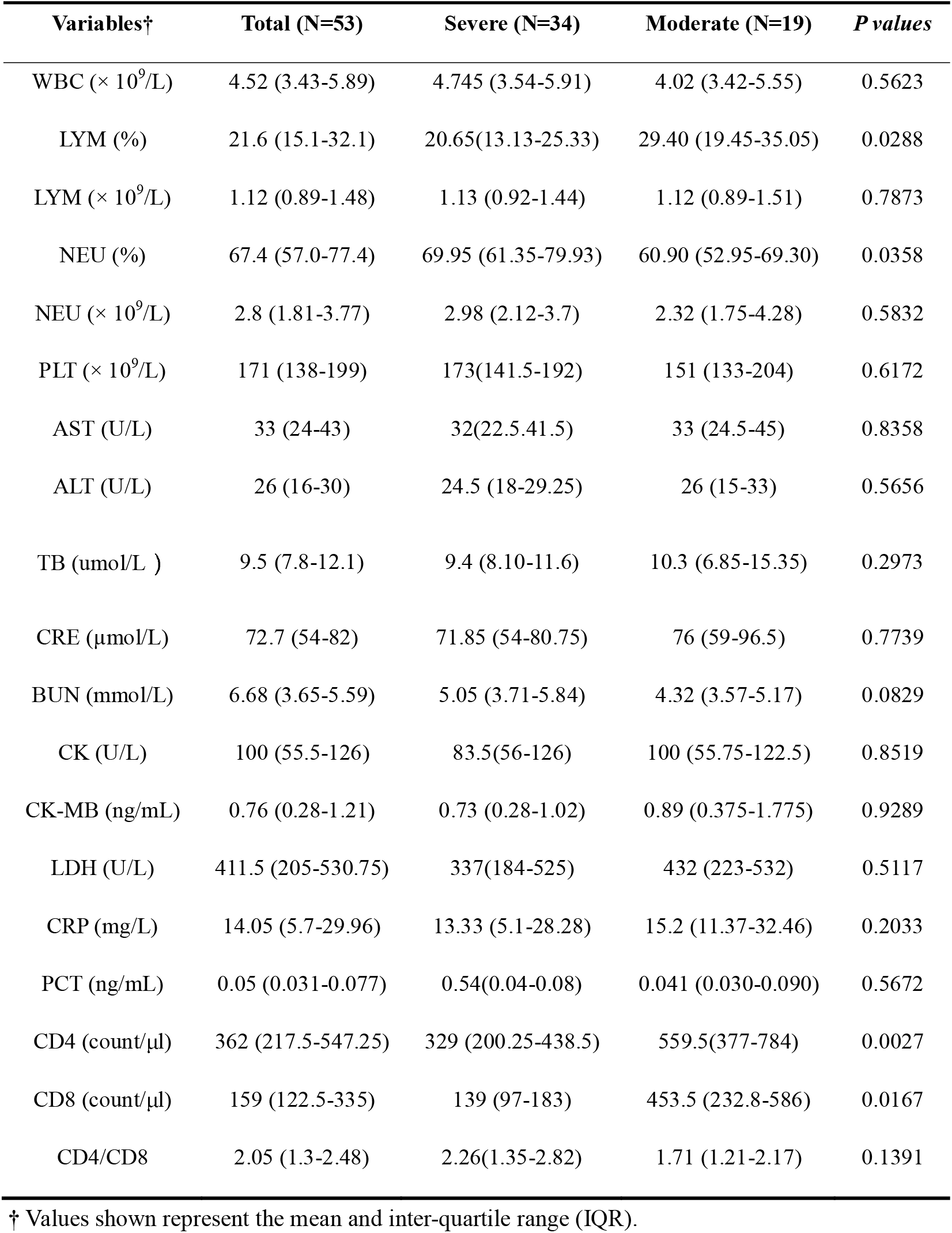
Laboratory results of COVID-19 patients in this study.

| Variables† | Total (N=53) | Severe (N=34) | Moderate (N=19) | *P values* |
| --- | --- | --- | --- | --- |
| WBC ($\times 10^9/L$) | 4.52 (3.43-5.89) | 4.745 (3.54-5.91) | 4.02 (3.42-5.55) | 0.5623 |
| LYM (%) | 21.6 (15.1-32.1) | 20.65(13.13-25.33) | 29.40 (19.45-35.05) | 0.0288 |
| LYM ($\times 10^9/L$) | 1.12 (0.89-1.48) | 1.13 (0.92-1.44) | 1.12 (0.89-1.51) | 0.7873 |
| NEU (%) | 67.4 (57.0-77.4) | 69.95 (61.35-79.93) | 60.90 (52.95-69.30) | 0.0358 |
| NEU ($\times 10^9/L$) | 2.8 (1.81-3.77) | 2.98 (2.12-3.7) | 2.32 (1.75-4.28) | 0.5832 |
| PLT ($\times 10^9/L$) | 171 (138-199) | 173(141.5-192) | 151 (133-204) | 0.6172 |
| AST (U/L) | 33 (24-43) | 32(22.5-41.5) | 33 (24.5-45) | 0.8358 |
| ALT (U/L) | 26 (16-30) | 24.5 (18-29.25) | 26 (15-33) | 0.5656 |
| TB (umol/L ) | 9.5 (7.8-12.1) | 9.4 (8.10-11.6) | 10.3 (6.85-15.35) | 0.2973 |
| CRE ($\mu\text{mol/L}$) | 72.7 (54-82) | 71.85 (54-80.75) | 76 (59-96.5) | 0.7739 |
| BUN (mmol/L) | 6.68 (3.65-5.59) | 5.05 (3.71-5.84) | 4.32 (3.57-5.17) | 0.0829 |
| CK (U/L) | 100 (55.5-126) | 83.5(56-126) | 100 (55.75-122.5) | 0.8519 |
| CK-MB (ng/mL) | 0.76 (0.28-1.21) | 0.73 (0.28-1.02) | 0.89 (0.375-1.775) | 0.9289 |
| LDH (U/L) | 411.5 (205-530.75) | 337(184-525) | 432 (223-532) | 0.5117 |
| CRP (mg/L) | 14.05 (5.7-29.96) | 13.33 (5.1-28.28) | 15.2 (11.37-32.46) | 0.2033 |
| PCT (ng/mL) | 0.05 (0.031-0.077) | 0.54(0.04-0.08) | 0.041 (0.030-0.090) | 0.5672 |
| CD4 (count/$\mu\text{l}$) | 362 (217.5-547.25) | 329 (200.25-438.5) | 559.5(377-784) | 0.0027 |
| CD8 (count/$\mu\text{l}$) | 159 (122.5-335) | 139 (97-183) | 453.5 (232.8-586) | 0.0167 |
| CD4/CD8 | 2.05 (1.3-2.48) | 2.26(1.35-2.82) | 1.71 (1.21-2.17) | 0.1391 |

† Values shown represent the mean and inter-quartile range (IQR).

Then the expression profile of cytokines/chemokines of COVID-19 cases upon admission was analyzed. Elevated concentrations of both pro- and anti-inflammatory cytokines were observed in these cases including IFN-γ (interferon, IFN), IL-1ra (interleukin, IL), IL-2ra, IL-6, IL-10, IL-18, HGF (hepatocyte growth factor), MCP-3 (monocyte chemotactic protein-3), MIG (monokine induced gamma interferon), M-CSF (macrophage colony stimulating factor), G-CSF (granulocyte colony-stimulating factor), MIG-1a (macrophage inflammatory protein 1 alpha) CTACK (cutaneous T-cell-attracting chemokine) and IP-10 (interferon gamma induced protein 10), when compared with healthy control, indicating that cytokine storm occurred following SARS-CoV-2 infection (Figure 1) as previously reported ^3,25^. Similar phenomenon was also observed in SARS-CoV ^17-21^ and MERS-CoV ^22-24^ infected patients, despite that the expression profiles were virus specific. Meanwhile, we compared the plasma cytokine/chemokine levels collected from severe and moderate cases at different days after illness onset (d.a.o). The results showed that most of the cytokines were comparable between the two groups and maintained high level even at the later stage of the disease (≥ 15 d.a.o). However, IP-10, MCP-3, IL-1ra and MIG were significantly higher in samples from severe cases in all the three groups of sample collection time, even though statistical significance was not observed in IL-1ra and MIG. Notably, IP-10 in the severe cases possessed a high and similar expression level at all groups of sample collection time. On the contrary, IP-10 in moderate cases was significantly lower and further decreased a lot at ≥ 15 d.a.o. Moreover, comparison of these fourteen cytokines between the severe and moderate cases showed significantly higher levels of IP-10, MCP-3 and IL-1ra in the severe cases (Figure 1). Spearman rank coefficient correlation analysis showed that IP-10, MCP-3 and IL-1ra expression levels were highly correlated with PaO_2_/FaO_2_ and Murray scores. Interestingly, the viral load (displayed as Ct values) showed obvious correlation with only IP-10, while not MCP-3 and IL-1ra, as well as the PaO_2_/FaO_2_ and Murray scores (Figure S2). Our previous study has found that the viral shedding in COVID-19 cases is complicated and the viral loads obtained from the regularly used upper respiratory specimens might not consistent with the real case, especially for the severe cases ^27^. Meanwhile, the main distribution of viruses in the severe and moderate cases were also different, which might influence the induction of cytokines as well. Indeed, studies have found that high viral loads in some types of viral infections correlated with severe diseases ^28-30^, while not for all of them ^31^. These results suggested that the production of specific cytokines while not the viral burden might play central in the pathogenesis COVID-19.

**Figure 1.**
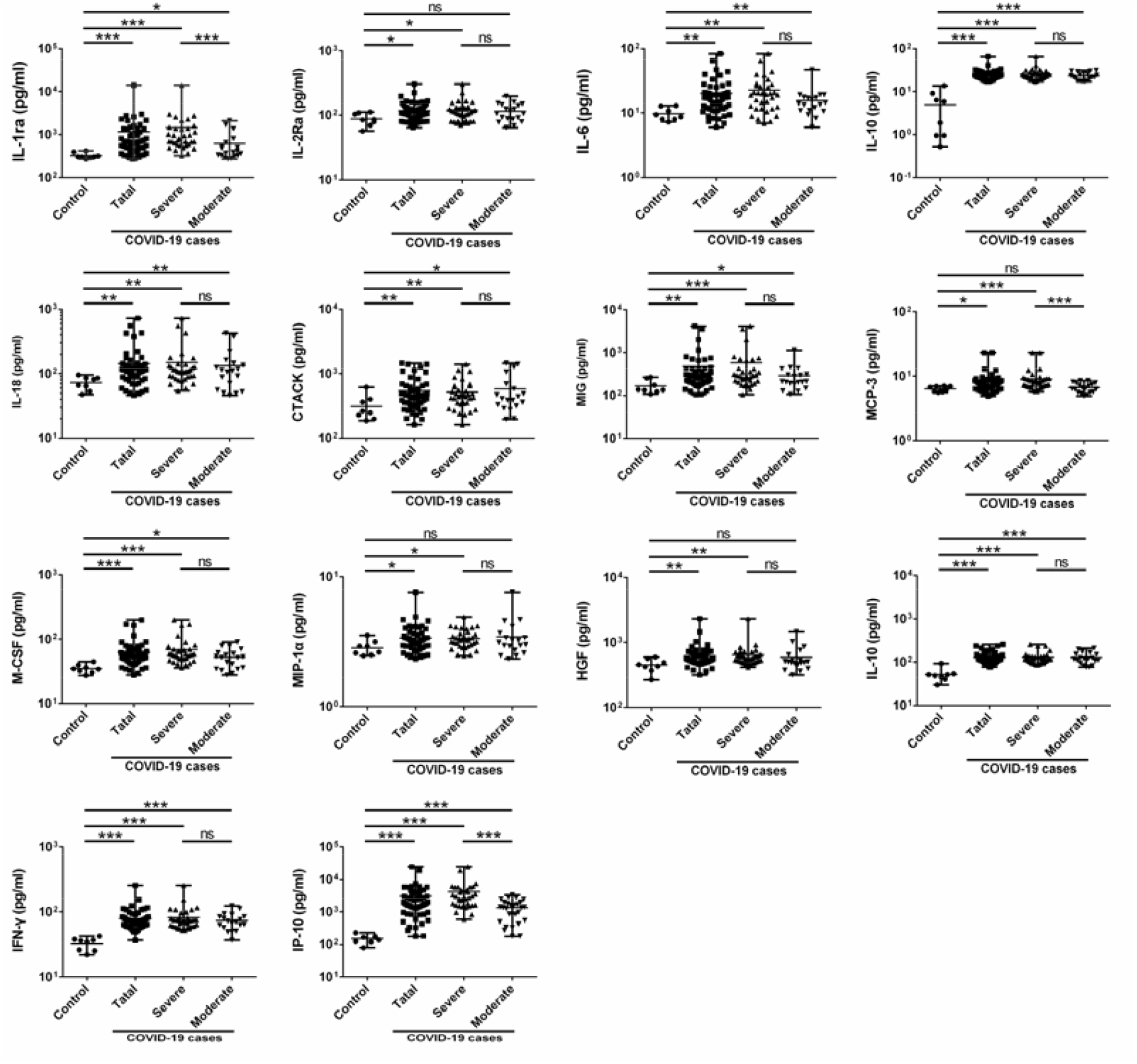
Comparison of serum cytokine/chemokine concentrations between healthy volunteers and COVID-19 cases. Samples from severe (N=34) and moderate (N=19) COVID-19 cases were collected at the earliest possible time-point after hospitalization for assays measuring the concentrations of 48 cytokines and chemokines, and healthy subjects (N=8) were involved as control. Values were graphed on a logarithmic scale and presented in units of pg/mL. *P values* between 0.01-0.05, 0.001-0.01 and 0.0001-0.001 were considered statistically significant (*), very significant (**) and extremely significant (***), respectively, whereas ns represents not significant.

We further analyzed whether these cytokines could be used as predictors for the disease severity and progression of COVID-19. Firstly, we calculated the receiver operating characteristic (ROC) of each single cytokine using the expression levels upon admission. Results showed that the area under the curve (AUC) of the ROC was 0.915, 0.765 and 0.792 for IP-10, MCP-3 and IL-1ra, respectively (Figure 3). Then we tested the different combination of these cytokines for the prediction of disease progression. Combinations of the three cytokines showed the highest AUC of 0.943, followed by the combination of IP-10 and MCP-3 (0.927) as well as the combination of IP-10 and IL-1ra (0.923) (Figure 3). Furthermore, we analyzed the serial change of the three cytokines in 14 severe cases. In the two fatal cases (cases 1 and 2), the expression levels were significantly high upon admission and maintained high levels during the disease progression. Meanwhile, five cases (cases 7∼9, 11 and 13) with continuously high levels of these cytokines, especially IP-10 and MCP-3, were still in critical condition. On the contrary, cases 3∼6, 10, 12 and 14 with lower levels or significantly decreased levels of these cytokines showed an obvious improvement of the disease, and became moderate cases (cases 4, 10, 12 and 14) or discharged from hospital (cases 3, 5, 6 and 10). These results indicated that these three cytokines could be considered as biomarkers to predict disease progression and outcome of SARS-CoV-2 infections.

**Figure 3.**
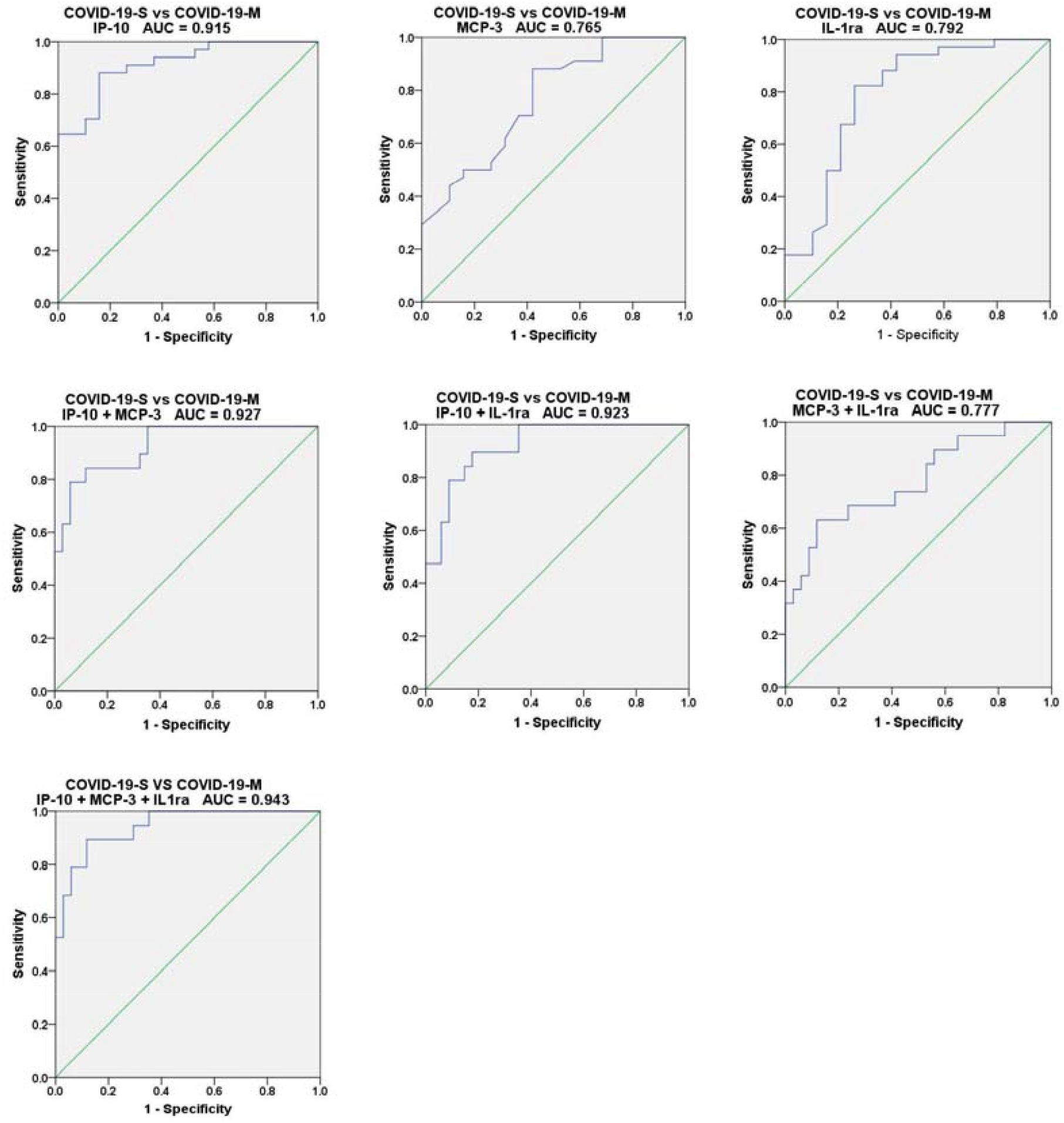
The ROC curve of plasma cytokine levels for patients with mild and severe SARS-CoV-2-infections. The receiver operating characteristic (ROC) curve was calculated using SPSS. The area under the curve (AUC) of ROC for IP-10, MCP-3 and IL-1ra wad estimated using the expression levels upon admission. The P values of all AUC for the three cytokines were less than 0.05.

Cytokine storm with uncontrolled proinflammatory responses induce significant immunopathology and severe disease outcomes during some viral infections, indicating an important role in the pathogenesis of these viruses ^2,32^. Accordingly, not only the pathogens but also the pathogen induced cytokine storm should be considered during the treatment. Therapy strategy with a combination of antimicrobial and immunotherapy may produce a more favorable outcome ^9^. Corticosteroids which could downregulate proinflammatory cytokine transcription and subsequently inhibit the cytokine storm ^33^, and the antiviral cytokine interferon-α were widely used during the treatment of COVID-19 ^8,34^. However, use of high-dose corticosteroids has been shown to be associated with an increase in mortality and significantly longer durations of viral shedding in H7N9 infected patients ^35^, which is of concern. The results of our study suggested a crucial role of IP-10 in the pathogenesis of SARS-CoV-2, which has also been shown to be associated with disease severity of H5N1, H1N1, SARS-CoV and MERS-CoV ^16,24,29,36^. A recent study has found that IP-10 (CXCL10)-CXCR3 signaling appears to be a critical factor for the exacerbation of the pathology of ARDS ^37^. Thus, modulators such as antibody targeting IP-10 might be a promising therapeutic strategy in the treatment of the acute phase of ARDS to ameliorates acute lung injury as shown in H1N1 mouse model ^37,38^.

In summary, we compared the differences of cytokines profiles between severe and moderate COVID-19 cases, and found that IP-10, MCP-3 and IL-1ra were significantly associated with disease severity and progression of COVID-19. Our results added to our understanding of the immunopathologic mechanisms of SARS-CoV-2 infection, and suggest a novel therapeutic strategy using modulators or antibodies against theses cytokines, especially IP-10.

## METHODS

### Patient information and data collection

Subjects presented in this study were hospitalized COVID-19 patients (N=53), among whom 34 were defined as severe cases according to China National Health Commission Guidelines for Diagnosis and Treatment of 2019-nCoV infection. Healthy controls (N=8) were also included. Clinical information and laboratory result were collected at the earliest time-point after hospitalization. The study protocol was approved by the Ethics Committees of Shenzhen Third People’s Hospital (SZTHEC2016001). Informed consents were obtained from all patients or patients family members. The study was conducted in accordance with the International Conference on Harmonisation Guidelines for Good Clinical Practice and the Declaration of Helsinki and institutional ethics guidelines.

### Quantitative reverse-transcriptase polymerase chain reaction (qRT-PCR)

qRT-PCR were done as previously reported ^26,34^. In brief, throat swabs, nasal swabs, sputum and BALF specimens were collected from laboratory-confirmed COVID-19 cases upon admission and thereafter. Viral RNAs were extracted from clinical specimens using the QIAamp RNA Viral Kit (Qiagen, Heiden, Germany). A China Food and Drug Administration (CDFA) approved commercially kit (GeneoDX Co., Shanghai, China) were used for the detection of SARS-CoV-2 specific RNAs. Samples with a cycle threshold (Ct) value ≤ 37.0 were considered putatively positive. Samples whose Ct was higher than 37 were re-tested and considered positive if Ct was ≥37 but ≤ 40 and negative if viral RNAs were undetectable on the second test.

### Disease severity classification

Disease severity classification and Murray Score calculation were evaluated as previously reported ^26^. Severity of 2019-nCoV infection was graded according to China National Health Commission Guidelines for Diagnosis and Treatment of SARS-CoV-2 infection. Laboratory confirmed patients with fever, respiratory manifestations and radiological findings indicative of pneumonia were considered moderate cases (COVID-19-M). Laboratory confirmed patients who met any of the following were considered to have severe COVID-19 (COVID-19-S): 1) respiratory distraction (respiration rate≥30/min; 2) resting oxygen saturation≤93%, or 3) arterial oxygen partial pressure (PaO_2_) / fraction of inspired oxygen (FiO_2_) ≤300 mmHg (1 mmHg = 0.133 kPa). Laboratory confirmed patients who had any of the following were considered in critical condition: 1) respiratory failure requiring mechanical ventilation, 2) shock, or 3) failure of other organs requiring intensive care unit (ICU).

### Quantification of hypoxia and Murray Score

The partial pressure of oxygen (PaO_2_) in arterial blood taken from the patients at various time-points after hospitalization is measured by the ABL90 blood gas analyzer (Radiometer). The fraction of inspired oxygen (FiO2) is calculated by the following formula: FiO_2_ = (21 + oxygen flow (in units of l/min) × 4) /100. The PaO_2_/FiO_2_ ratio (in units of mmHg) is calculated by dividing the PaO_2_ value with the FiO_2_ value. A PaO_2_/FiO_2_ ratio less than or equal to 100 mmHg is considered one of the criteria for severe acute respiratory distress syndrome (ARDS). Murray Score were calculated as reported ^39^.

### Cytokine and chemokine measurements

The plasma of patients with laboratory-confirmed COVID-19 cases (N=53) were collected at the earliest possible time-point after hospitalization and thereafter. The plasma of healthy subjects (N=8) were included as the negative control group. The concentrations of 48 cytokines and chemokines were measured using Bio-Plex Pro Human Cytokine Screening Panel (Bio-Rad) as previously reported ^15,26^.

### Statistical analysis

Fisher exact test was used to compare the indicated rates between the moderate and severe cases. Mann-Whitney U test was used to compare plasma cytokine levels between two groups. The Spearman rank correlation coefficient was used for linear correlation analysis between the expression level of plasma cytokine and PaO2/FiO2 ratio, Murray score as well as viral loads of COVID-19 cases. The area under the receiver operating characteristic (ROC) curve (AUC) of plasma cytokine levels was estimated for the moderate and severe cases. All statistical tests were calculated using SPSS 20.0 for Windows (IBM, Chicago, USA). *P value* less than 0.05 was considered statistically significant.

## Data Availability

All data generated or used during the study have been presented in the submitted article.

## Acknowledgement

This work was supported by the National Natural Science Foundation of China (Grant 81802004) and the National Science and Technology Major Project (Grants 2018ZX10711001, 2017ZX10204401 and 2017ZX10103011), Shenzhen Science and Technology Research and Development Project (Grants JCYJ20180228162201541, 202002073000001) Sanming Project of Medicine in Shenzhen (SZSM201412003, SZSM201512005).

## Author contributions

Y. Liu, L. Liu, Z. Zhang and Y. Yang conceived and guided the project. Y. Yang, C. Shen, M. Yang carried out the experiments and analyzed the data. J. Li, J. Yuan, F. Wang, Z. Wang, M. Cao, W. Wu, R. Zou, Y. Li, L. Xing, L. Peng, J. Wei, H. Zheng, Z. Xu, and H. Wang collected clinical samples and the data. Y. Yang wrote the manuscript. All the authors have read and approved the manuscript.

## Competing interests

The authors declare no competing interests.

**Figure 4.**
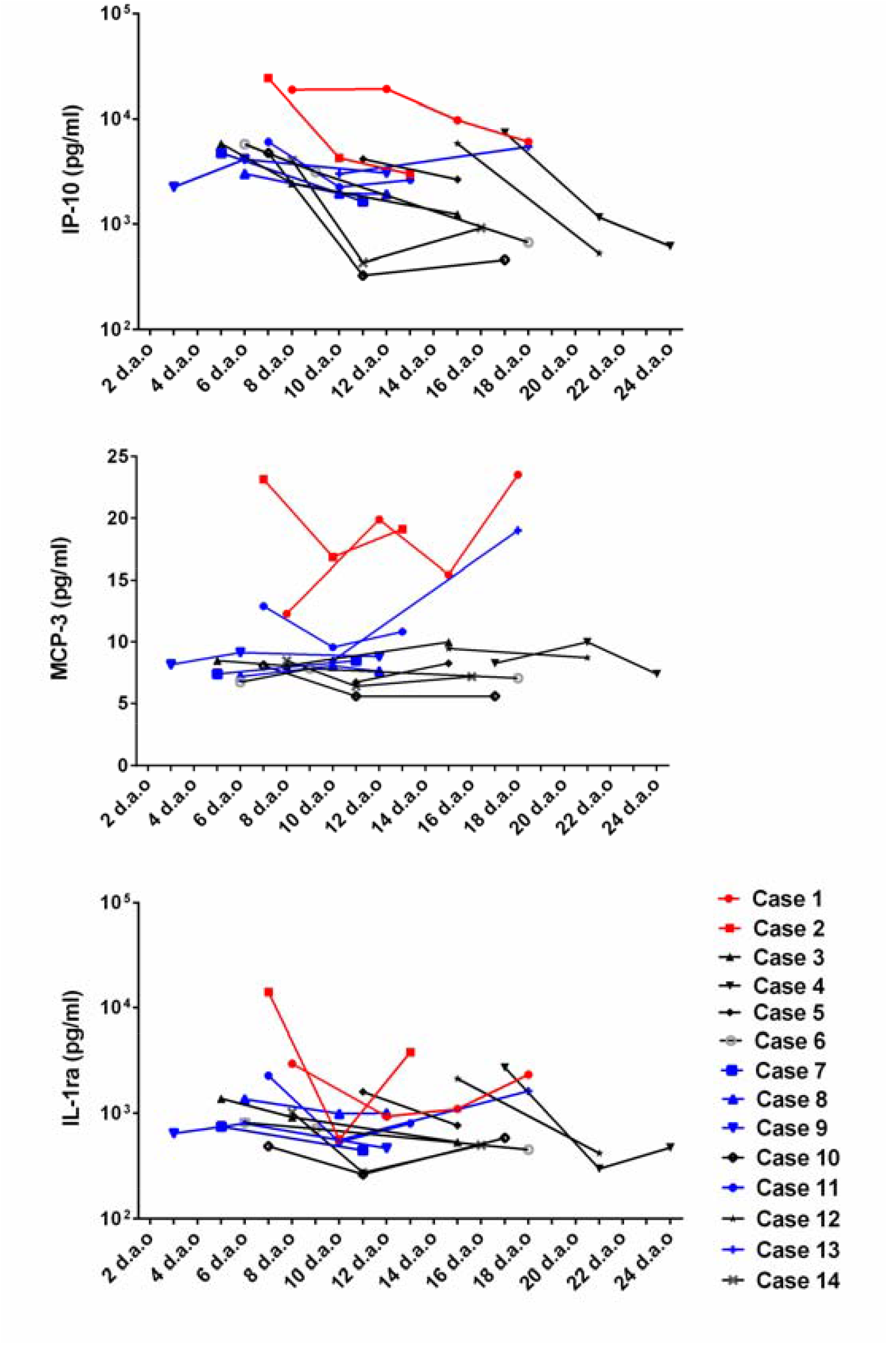
Dynamic changes of IP-10, MCP-10 and IL-1ra in 14 severe cases of COVID-19 with different outcome and progression. The expression levels measured at the indicated time-points of 14 severe cases were shown. The fatal cases were marked in red. Cases in critical condition were marked in blue, and the others were cases with moderate disease or discharged at present.

**Figure 2.**
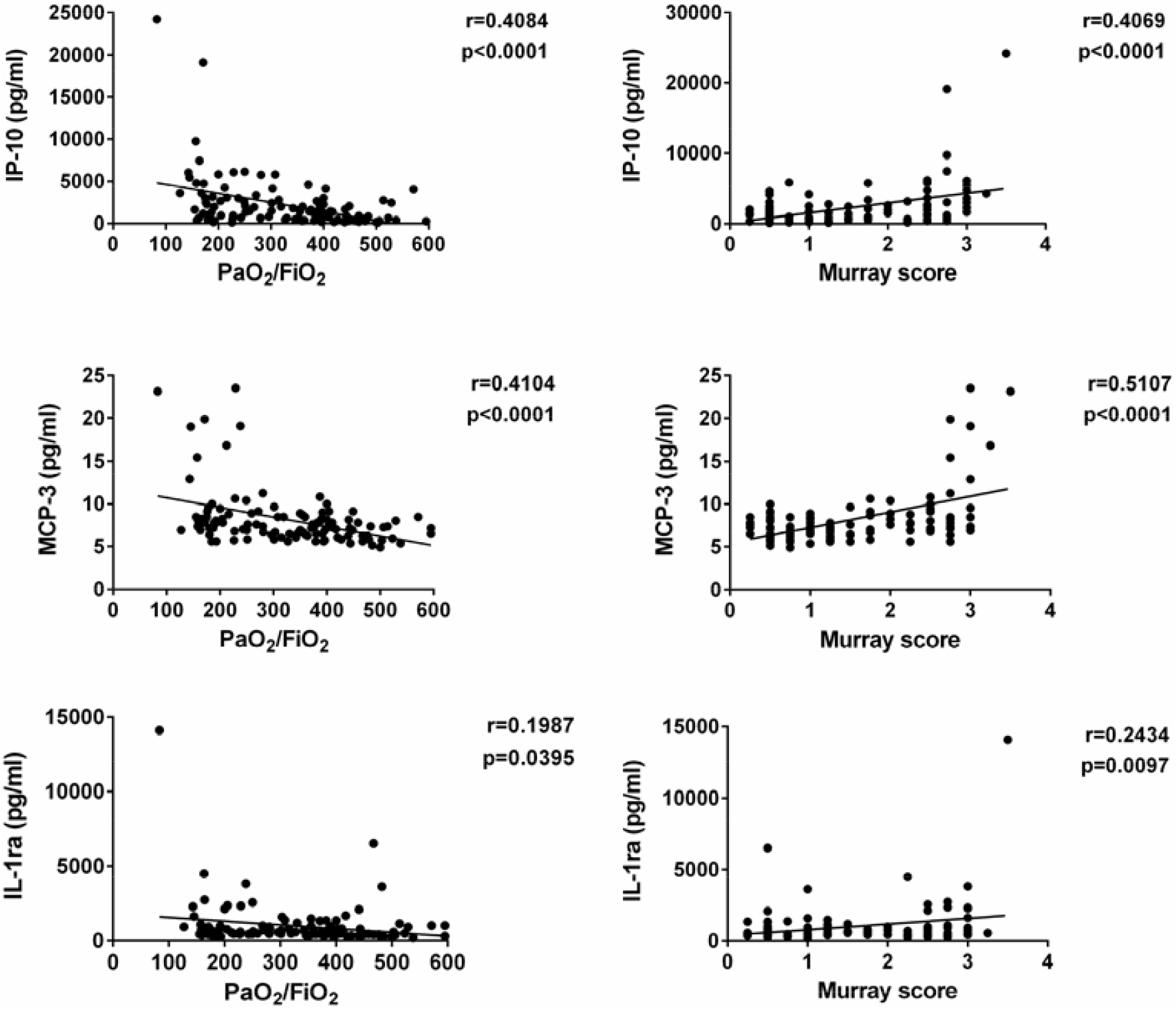
IP-10, MCP-3 and IL-1ra expression levels were highly correlated with PaO_2_/FiO_2_ and Murray Score. The expression levels of IP-10, MCP-3 and IL-1ra measured from plasma samples collected upon admission and thereafter and the corresponding PaO2/FiO2 and Murray Score at the same day were analyzed using spearman rank correlation analysis. The *r values* and *p values* were shown.

**Figure S1.**
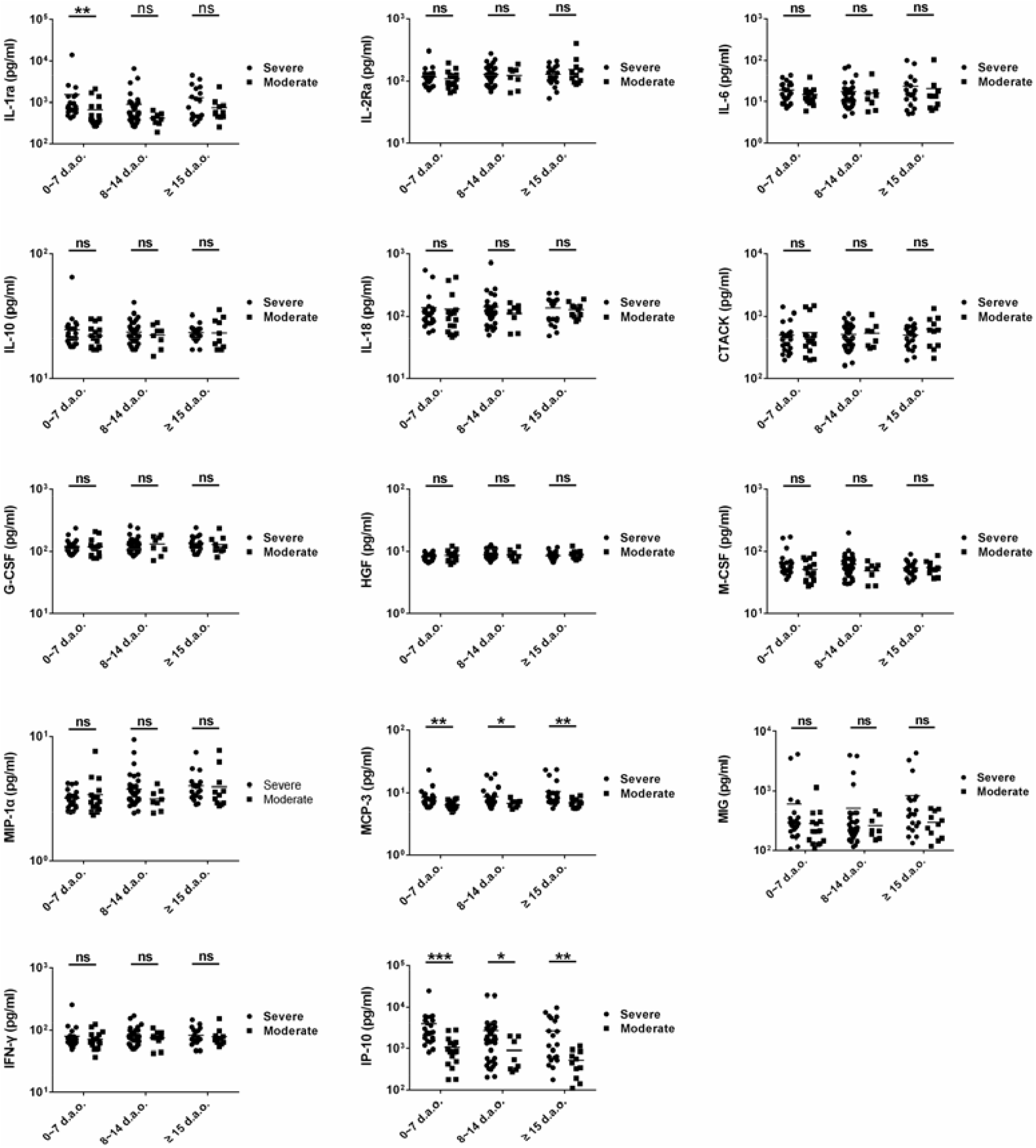
Comparison of the significantly elevated cytokines measured at different days post illness onset between severe and moderate COVID-19 cases. The expression levels of 14 significantly elevated cytokines measured at different days post illness onset were shown and compared between the severe and moderate COVID-19 cases. *P values* between 0.01-0.05, 0.001-0.01 and 0.0001-0.001 were considered statistically significant (*), very significant (**) and extremely significant (***), respectively, whereas ns represents not significant.

**Figure S2.**
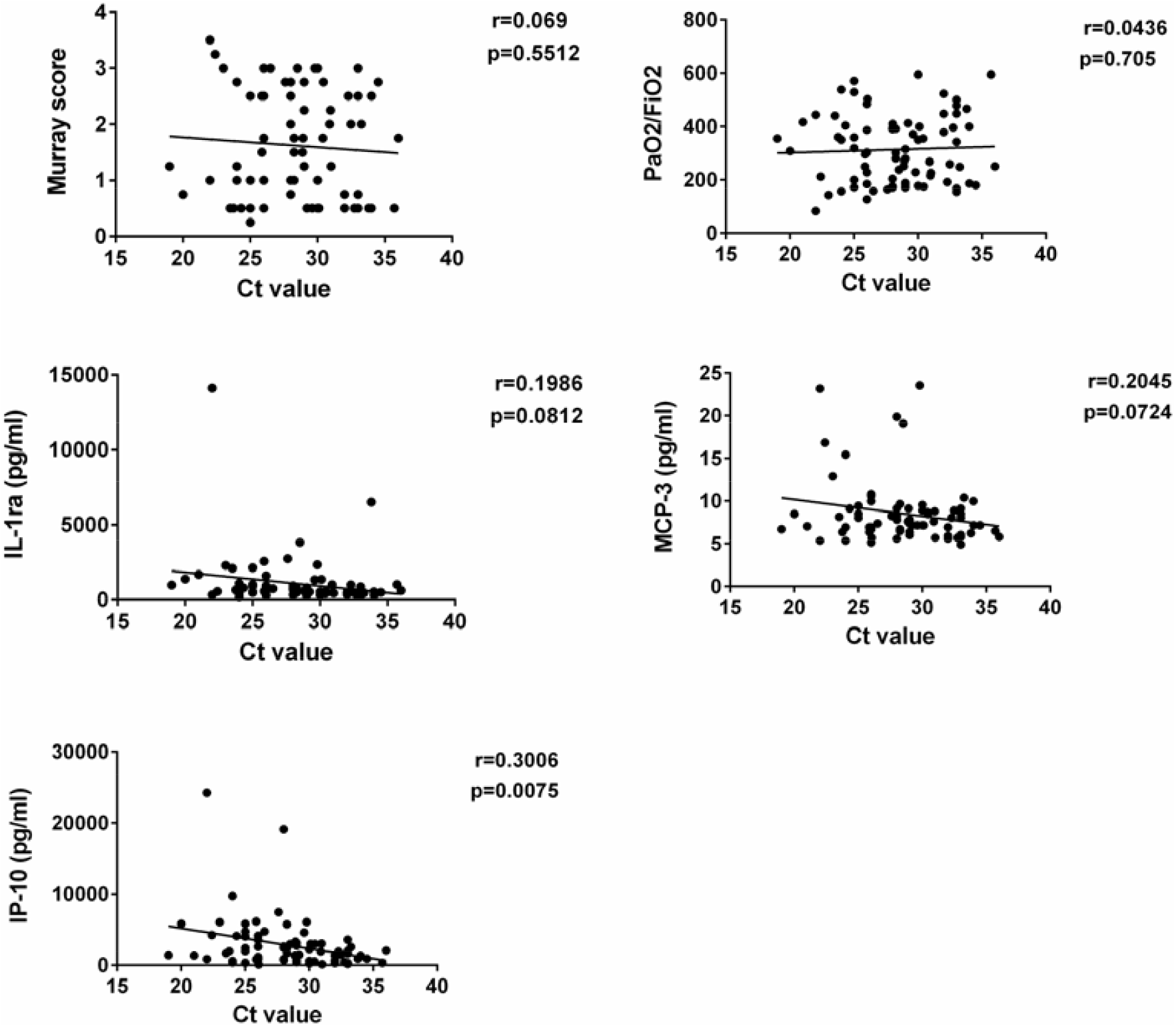
Correlations between viral load and the cytokines of IP-10, MCP-3 and IL-1ra, as well as PaO2/FiO2 and Murray Score. The correlations between viral load and the cytokines of IP-10, MCP-3 and IL-1ra, as well as PaO2/FiO2 and Murray Score were analyzed using spearman rank correlation analysis. The *r values* and *p values* were shown.The viral loads were indicated as Ct values, and lower Ct values represented high viral load.

**Table S1.**
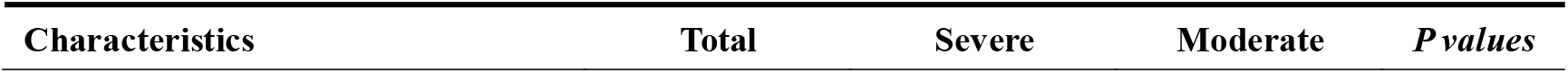
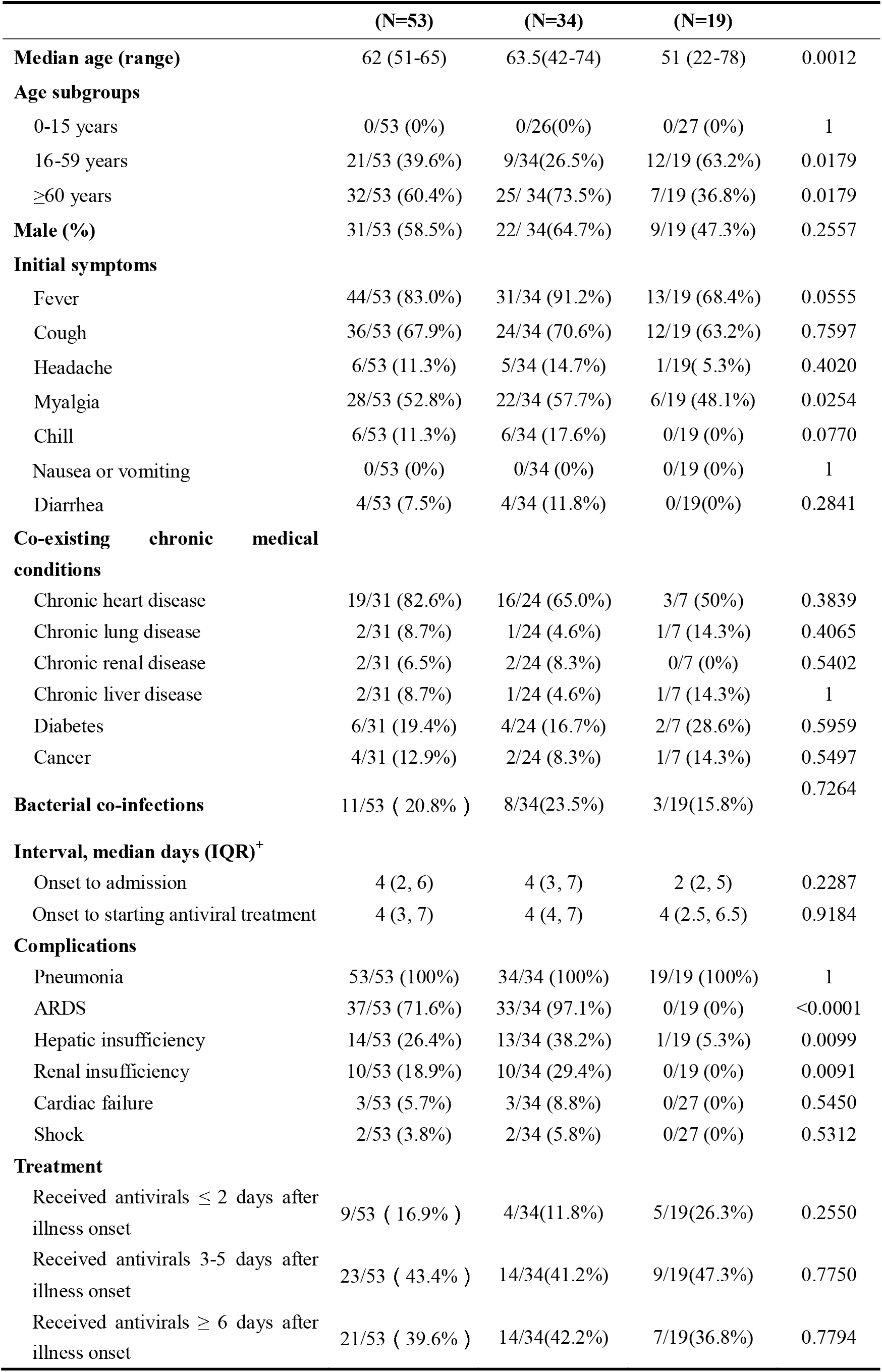
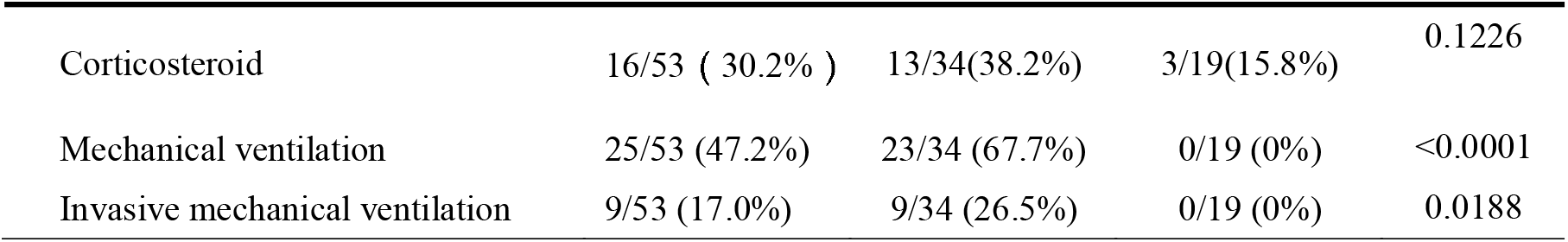
Epidemiological and clinical features of COVID-19 patients in this study. Characteristics Total Severe Moderate *P values*

|  | (N=53) | (N=34) | (N=19) |  |
| --- | --- | --- | --- | --- |
| **Median age (range)** | 62 (51-65) | 63.5(42-74) | 51 (22-78) | 0.0012 |
| **Age subgroups** |  |  |  |  |
| 0-15 years | 0/53 (0%) | 0/26(0%) | 0/27 (0%) | 1 |
| 16-59 years | 21/53 (39.6%) | 9/34(26.5%) | 12/19 (63.2%) | 0.0179 |
| ≥60 years | 32/53 (60.4%) | 25/ 34(73.5%) | 7/19 (36.8%) | 0.0179 |
| **Male (%)** | 31/53 (58.5%) | 22/ 34(64.7%) | 9/19 (47.3%) | 0.2557 |
| **Initial symptoms** |  |  |  |  |
| Fever | 44/53 (83.0%) | 31/34 (91.2%) | 13/19 (68.4%) | 0.0555 |
| Cough | 36/53 (67.9%) | 24/34 (70.6%) | 12/19 (63.2%) | 0.7597 |
| Headache | 6/53 (11.3%) | 5/34 (14.7%) | 1/19( 5.3%) | 0.4020 |
| Myalgia | 28/53 (52.8%) | 22/34 (57.7%) | 6/19 (48.1%) | 0.0254 |
| Chill | 6/53 (11.3%) | 6/34 (17.6%) | 0/19 (0%) | 0.0770 |
| Nausea or vomiting | 0/53 (0%) | 0/34 (0%) | 0/19 (0%) | 1 |
| Diarrhea | 4/53 (7.5%) | 4/34 (11.8%) | 0/19(0%) | 0.2841 |
| **Co-existing chronic medical conditions** |  |  |  |  |
| Chronic heart disease | 19/31 (82.6%) | 16/24 (65.0%) | 3/7 (50%) | 0.3839 |
| Chronic lung disease | 2/31 (8.7%) | 1/24 (4.6%) | 1/7 (14.3%) | 0.4065 |
| Chronic renal disease | 2/31 (6.5%) | 2/24 (8.3%) | 0/7 (0%) | 0.5402 |
| Chronic liver disease | 2/31 (8.7%) | 1/24 (4.6%) | 1/7 (14.3%) | 1 |
| Diabetes | 6/31 (19.4%) | 4/24 (16.7%) | 2/7 (28.6%) | 0.5959 |
| Cancer | 4/31 (12.9%) | 2/24 (8.3%) | 1/7 (14.3%) | 0.5497 |
| **Bacterial co-infections** | 11/53 ( 20.8% ) | 8/34(23.5%) | 3/19(15.8%) | 0.7264 |
| **Interval, median days (IQR)<sup>+</sup>** |  |  |  |  |
| Onset to admission | 4 (2, 6) | 4 (3, 7) | 2 (2, 5) | 0.2287 |
| Onset to starting antiviral treatment | 4 (3, 7) | 4 (4, 7) | 4 (2.5, 6.5) | 0.9184 |
| **Complications** |  |  |  |  |
| Pneumonia | 53/53 (100%) | 34/34 (100%) | 19/19 (100%) | 1 |
| ARDS | 37/53 (71.6%) | 33/34 (97.1%) | 0/19 (0%) | <0.0001 |
| Hepatic insufficiency | 14/53 (26.4%) | 13/34 (38.2%) | 1/19 (5.3%) | 0.0099 |
| Renal insufficiency | 10/53 (18.9%) | 10/34 (29.4%) | 0/19 (0%) | 0.0091 |
| Cardiac failure | 3/53 (5.7%) | 3/34 (8.8%) | 0/27 (0%) | 0.5450 |
| Shock | 2/53 (3.8%) | 2/34 (5.8%) | 0/27 (0%) | 0.5312 |
| **Treatment** |  |  |  |  |
| Received antivirals ≤ 2 days after illness onset | 9/53 ( 16.9% ) | 4/34(11.8%) | 5/19(26.3%) | 0.2550 |
| Received antivirals 3-5 days after illness onset | 23/53 ( 43.4% ) | 14/34(41.2%) | 9/19(47.3%) | 0.7750 |
| Received antivirals ≥ 6 days after illness onset | 21/53 ( 39.6% ) | 14/34(42.2%) | 7/19(36.8%) | 0.7794 |
| Corticosteroid | 16/53 ( 30.2% ) | 13/34(38.2%) | 3/19(15.8%) | 0.1226 |
| Mechanical ventilation | 25/53 (47.2%) | 23/34 (67.7%) | 0/19 (0%) | <0.0001 |
| Invasive mechanical ventilation | 9/53 (17.0%) | 9/34 (26.5%) | 0/19 (0%) | 0.0188 |

